# Liver-Quant: Feature-Based Image Analysis Toolkit for Automatic Quantification of Metabolic Dysfunction-Associated Steatotic Liver Disease

**DOI:** 10.1101/2024.05.21.24305727

**Authors:** Mohsen Farzi, Clare McGenity, Alyn Cratchley, Leo Leplat, Peter Bankhead, Alexander Wright, Darren Treanor

**Affiliations:** Department of Histopathology, Leeds Teaching Hospitals NHS Trust, Leeds, UK; University of Leeds, Leeds, UK; Department of Clinical Pathology & Department of Clinical and Experimental Medicine, Linköping University, Linköping, Sweden; Centre for Medical Image Science and Visualization (CMIV), Linköping University, Linköping, Sweden; Centre for Genomic & Experimental Medicine, Institute of Genetics and Cancer, The University of Edinburgh, Edinburgh, UK; Edinburgh Pathology and CRUK Scotland Centre, Institute of Genetics and Cancer, The University of Edinburgh, Edinburgh, UK

## Abstract

**Introduction:** The histological assessment of liver biopsies by pathologists serves as the gold standard for diagnosing metabolic dysfunction-associated steatotic liver disease (MASLD) and staging disease progression. Various machine learning and image analysis tools have been reported to automate the quantification of fatty liver and enhance patient risk stratification. However, the current software is either not open-source or not directly applicable to the whole slide images (WSIs).

**Methods:** In this paper, we introduce “*Liver-Quant*,” an open-source Python package designed for quantifying fat and fibrosis in liver WSIs. Employing colour and morphological features, Liver-Quant measures the Steatosis Proportionate Area (SPA) and Collagen Proportionate Area (CPA). The method’s accuracy and robustness were evaluated using an internal dataset of 424 WSIs from adult patients collected retrospectively from the archives at Leeds Teaching Hospitals NHS Trust between 2016 and 2022 and an external public dataset of 109 WSIs. For each slide, semi-quantitative scores were automatically extracted from free-text pathological reports. Furthermore, we investigated the impact of three different staining dyes including Van Gieson (VG), Picro Sirius Red (PSR), and Masson’s Trichrome (MTC) on fibrosis quantification.

**Results:** The Spearman rank coefficient (ρ) was calculated to assess the correlation between the computed SPA/CPA values and the semi-quantitative pathologist scores. For steatosis quantification, we observed a substantial correlation (ρ=0.92), while fibrosis quantification exhibited a moderate correlation with human scores (ρ=0.51). To assess stain variation on CPA measurement, we collected N=18 cases and applied the three stains. Employing stain normalisation, an excellent agreement was observed in CPA measurements among the three stains using Bland-Altman plots. However, without stain normalisation, PSR emerged as the most effective dye due to its enhanced contrast in the Hue channel, displaying a strong correlation with human scores (ρ=0.9), followed by VG (ρ=0.8) and MTC (ρ=0.59). Additionally, we explored the impact of the apparent magnification on SPA and CPA. High resolution images collected at 0.25 microns per pixel (MPP) [apparent magnification = 40x] or 0.50 MPP [apparent magnification = 20x] were found to be essential for accurate SPA measurement, whereas for CPA measurement, low resolution images collected at 10 MPP [apparent magnification = 1x] were sufficient.

**Conclusion:** The Liver-Quant package offers an open-source solution for rapid and precise liver quantification in WSIs applicable to multiple histological stains.

## 1. Introduction

Non-alcoholic fatty liver disease (NAFLD) or metabolic dysfunction-associated steatotic liver disease (MASLD) is characterised by the excessive accumulation of fat in the liver, independent of significant alcohol use or other known liver diseases [1], [2]. MASLD represents a spectrum of disease; the majority of patients present with a mild disease termed simple steatosis, which typically does not progress further. However, a subset of patients may develop a more severe form, known as non-alcoholic steatohepatitis (NASH), which can progress to varying degrees of fibrosis, cirrhosis, and increase the risk of developing hepatocellular carcinoma (HCC) [1]. The global prevalence of NASH is rapidly growing and causes a considerable burden on healthcare systems [3], [4].

Histological assessment of liver biopsies by a pathologist is still considered the *gold standard* for diagnosing NASH and staging the disease [5]. The Non-alcoholic fatty liver disease Activity Score (NAS) or NASH Clinical Research Network (NASH CRN) score developed by Kleiner *et al.* [6] is commonly used for the diagnosis and staging of NASH. NAS is defined as the sum of the scores for three histological features - steatosis (0-3), lobular inflammation (0-3), and ballooning (0-2); resulting in a score ranging from 0 to 8 [6]. In the Kleiner system, steatosis in liver biopsies is reported semi-quantitatively using a four-graded scale: 0 (none), 1 (mild), 2 (moderate), or 3 (severe). Scores 0 to 3 are considered to correspond to fat percentages of less than 5%, 5-33%, 33-66%, and over 66% in the hepatocytes, respectively [6]. The fibrosis stage is also determined using a semi-quantitative score: 0 (no evidence of fibrotic changes or scarring), 1 (mild fibrosis confined to the area around portal tracts or sinusoids), 2 (perisinusoidal and portal/periportal fibrosis), 3 (bridging fibrosis), and 4 (cirrhosis).

Employing a subjective semi-quantitative scoring system by pathologists has four main drawbacks: a) pathologists tend to significantly overestimate the amount of fat in the liver [7]; b) subjective scores have only moderate reproducibility in terms of inter- and intra-observer variability [8]; c) ordinal classification has a limited ability to describe disease severity as a continuum, making borderline cases challenging; and d) ordinal classification has limited sensitivity to reflect disease progression over time and in response to treatment [9].

During the last decade, advances in technology have facilitated the wider adoption of digital pathology. In digital pathology, high-resolution whole slide images (WSIs) of the entire tissue section are generated using specialized scanning devices, usually at 0.25 or 0.50 microns per pixel (MPP) corresponding to apparent magnification of 40x or 20x, respectively [10]. Digital pathology opens opportunities to employ image analysis algorithms and artificial intelligence techniques to aid pathologists in diagnosing and predicting outcomes in liver histopathology [11].

For routine assessment of liver biopsies with steatosis, haematoxylin and eosin (H&E) are the preferred dyes to stain the tissue. The haematoxylin stains cell nuclei a purplish blue, and the eosin stains cytoplasm and the extra-cellular matrix pink. Fat is dissolved during the tissue processing procedure, and thus, fat globules appear as empty spaces within the tissue represented by a white colour (c.f. Figure 1) [12]. Various classical image analysis methods have been reported to quantify fat in the liver [13], [14], [15]. The overall procedure typically includes segmenting of the background and estimating of the tissue area, splitting the image into overlapping tiles, segmenting the white regions inside the tissue, differentiating between fat globules and other white regions, and finally computing the Steatosis Proportionate Area (SPA). To distinguish between fat globules and other white structures like central veins, portal veins, portal arteries, sinusoids, and bile ducts, both unsupervised and supervised methods have been proposed. Unsupervised learning methods rely on the assumption that fat globules often exhibit a mostly circular shape. For instance, a study by Forlano et al.[15] employed circular masks of different sizes in an iterative procedure to remove non-fat globules. Supervised learning methods involve data annotation such as polygons indicating the fat globules [9] or patch-level scores [16]. Classification can be approached as a two-class problem [13], distinguishing between fat and non-fat regions, or as a multi-class problem [14]. Traditional machine learning algorithms such as random forest [17] or support vector machines (SVMs) [18] have also been used for classification. Overall, these methods performed well in identifying fat globules with sensitivity and specificity reported to be above 90% [13], [14]. However, their efficacy in measuring SPA and its correlation with Kleiner scores has not been investigated. Deep learning techniques, including convolutional neural networks (CNNs), have also shown promising results in automatic fat quantification in MASLD [9], [16], [19], [20], [21], [22].

**Figure 1.**
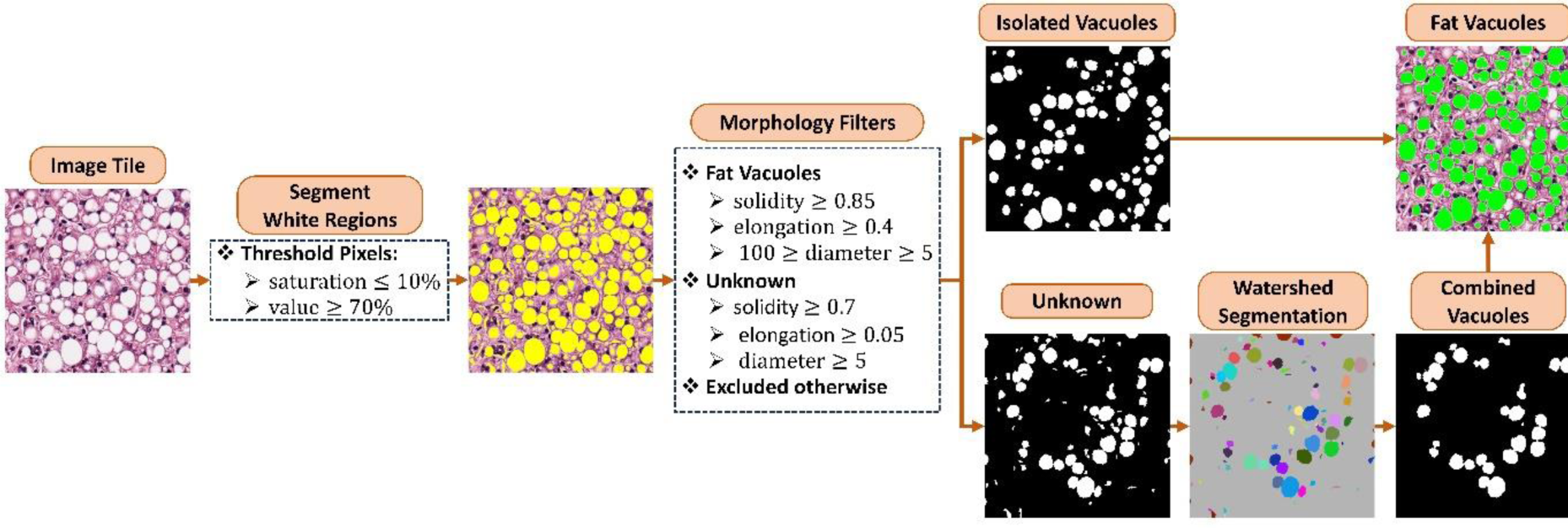
Fat Quantification Workflow. White regions are identified through pixel thresholding in the HSV space, with the resulting white space highlighted in yellow. Next, three shape descriptors are computed for each isolated region, namely, solidity (the ratio of the blob area to its convex hull area), elongation (the ratio of the long to the short axis), and the diameter of the enclosing circle. Based on these shape descriptors, the blobs are classified into three categories: isolated fat globules, combined blobs potentially containing overlapping fat globules (referred to as unknown), and other structures such as sinusoids, vessels, and portal tracts. The unknown white regions are further segmented into smaller regions using watershed segmentation. Morphological filters are then applied to identify isolated fat globules. Finally, the resulting fat globules, both isolated and combined, are marked in green.

To quantify fibrosis, special histochemical staining techniques are used to highlight collagen fibres in the tissue sample. Picro Sirius red (PSR), Masson’s Trichrome (MTC), and Van Gieson (VG) staining are three widely used methods to visualize and quantify the collagen content within liver tissue (Figure 2). PSR staining highlights collagen fibres and helps differentiate between mature (thick, red-orange staining) and immature (thin, green staining) collagen. MTC staining highlights collagen fibres as blue, nuclei as dark blue or black, and cytoplasm and muscle fibres as red. VG staining highlights collagen fibres red or pink, while other tissue components such as muscle fibres, cytoplasm, and nuclei are stained yellow or brown. The use of these stains varies in different laboratories; PSR is the preferred histochemical method for automatic computer-based fibrosis evaluation due to its excellent binding affinity for collagen fibres and enhanced contrast [23], [24], whereas MTC and VG are more widely used to stain liver biopsies in clinical practice [25], [26].

**Figure 2.**
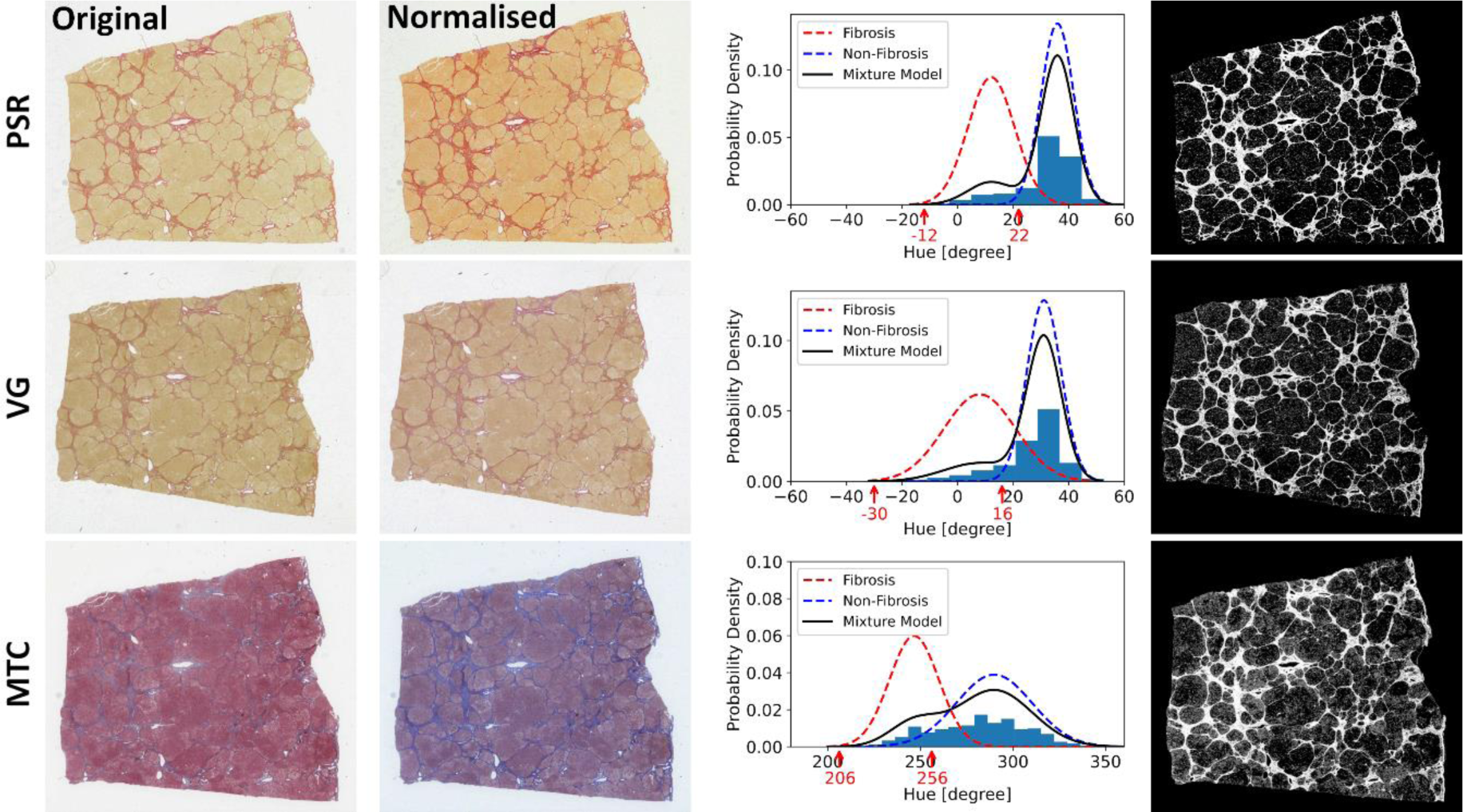
Fibrosis quantification. A sample biopsy with a fibrosis stage of 3 is shown in the first column stained with PSR, VG, and MTC, respectively. Prior to colour segmentation, stain normalisation is essential to remove differences in colour profiles and standardise the stain appearance [34]. Next, a Gaussian mixture model is fitted to the histogram of hue values and optimal thresholds (shown with red arrows) are selected. Binary masks are generated using pixel thresholding in the HSV-space. The estimated collagen proportionate area (CPA) was 16.7%, 14.0%, and 18.5% for PSR, VG, and MTC respectively.

Machine learning tools to quantify fibrosis are broadly divided into two groups[27]. In the first group, Collagen Proportionate Area (CPA) is measured to quantify the extent of liver fibrosis [28], [29], [30], [31]. Two main approaches have been reported in the literature to estimate CPA (Table 1): colour segmentation and deep learning methods. In colour segmentation, different techniques are employed such as thresholding in the RGB space [29], thresholding in the HSV space, and colour deconvolution followed by thresholding. On the other hand, deep learning methods utilize a U-Net architecture, which is commonly used for image segmentation tasks. In this second group, rather than estimating an intermediate biomarker like CPA, scores similar to those reported by pathologists in the Kleiner system are directly predicted. Estimating pathologist-like scores for the quantification of fibrosis has also been addressed using traditional feature learning and classification algorithms like SVM and recently deep learning frameworks.

**Table 1:** Image analysis methods to quantitate steatosis and fibrosis ^a^.

|  | Method Summary | Stain | N | Fibrosis | Steatosis |
| --- | --- | --- | --- | --- | --- |
| Masseroli et al. (2000) [28] | Automatic Maximum Likelihood thresholding by converting RGB image into grey-scale | PSR | 30 | $\rho = 0.72-0.83$ | - |
| Calvaruso et al. (2009) [29] | semi-automatic thresholding in RGB space using Zeiss KS300 image analysis software | PSR | 244 | $\rho = 0.6$ | - |
| Vanderbeck et al. (2013) [14] | White region classification using Support Vector Machine (SVM) trained on statistical features extracted from N=1969 white regions | H&E | 47 | - | $\rho = 0.88^b$ |
| Buzzetti et al. (2019) [30] | Semi-automatic thresholding in RGB space using Zeiss Axiovision (version 4.8.2.); confounding artefacts such as major blood vessels and liver capsules were manually removed. | PSR | 437 | - | - |
| Forlano et al. (2020) [15] | Steatosis quantification: use circular masks of different sizes in an iterative procedure to remove non-fat globules.<br>Fibrosis quantification: supervised liver tissue detection followed by K-means clustering using RGB values to detect fibrotic tissue | H&E<br>PSR | Train: 100<br>Test: 146 | $\rho = 0.57$ | $\rho = 0.66$ |
| Gawrieh et al. (2020) [31] | Automatic; universal RGB thresholding: normalised blue channel in [0.6863-0.9411], red and green channels in [0.2941-0.8235]; collagen area is the sum of detected regions weighted by the blue content. | MTC | 18 | $\rho = 0.77-0.93$ | - |
| Taylor et al. (2021) [9] | Network topology: 8-12 blocks of compound layers Inspired by residual networks and inception<br>Annotation: pixel-level polygons (n>65,424) | H&E<br>MTC | Train: 1255<br>Test: 5139 | $\rho = 0.63$ | $\rho = 0.6$ |
| Heinemann et al. (2022) [16] | Network topology: Inception-V3<br>Annotation: image tiles with Kleiner score | MTC | Train: 296<br>Test: 171 | $\kappa = 0.62$ | $\kappa = 0.66$ |

<sup>a</sup> $\rho$ = Spearman Rank Correlation Index and $\kappa$ = quadratic weighted Cohen's $\kappa$ on the test set
<sup>b</sup> The correlation index is computed using figures provided in the manuscript.

Several machine-learning tools have been reported to quantify fat and fibrosis in the liver [27], [32]. Despite their merits, unmet challenges still exist. First, there are very few open-source tools available for automatic quantitative analysis of fatty liver disease in WSIs [16]. Second, there are significant inconsistencies in calculated CPA and fat percentage by automated machine-learning algorithms. No standard automatic method currently exists with validation on benchmark datasets. Finally, staining standardization and colour adjustment may be necessary if machine learning-assisted scoring of stains is to be widely used.

The objective of this study was to leverage feature-based image analysis to quantitate steatosis and fibrosis using WSIs in MASLD. To this end, this paper presents an open-source Python-based software solution called ‘*Liver-Quant*’. The proposed algorithm is based on intuitive colour-based and morphological features to identify fat globules and scares in the tissue without the need for any manual annotations. To evaluate the accuracy and robustness of our method, we conducted an extensive analysis of N=424 WSIs stained with various dyes, including Haematoxylin and Eosin (H&E), Picro Sirius Red (PSR), Masson’s Trichrome (MTC), and Van Gieson (VG). Additionally, we explored the impact of magnification levels on the performance of quantification algorithms, optimizing for both accuracy and run time.

## 2. Materials and Methods

### 2.1. Study Design

The aim of this study is to use colour-based and morphological features to develop an open-source software solution to quantitate steatosis and fibrosis in liver WSIs. Specifically, we quantitate two key metrics: SPA and CPA. Additionally, we assess the accuracy and robustness of these metrics under varying staining techniques and scanning magnifications. This study serves as an exploratory investigation into the correlation patterns between semi-quantitative pathological staging of MASLD and the two quantitative metrics, SPA and CPA. We achieve this by analysing retrospective data obtained from human medical liver biopsies from two different datasets. Through this exploration, we aim to uncover insights that may inform further research in liver disease diagnosis and management.

### 2.2. Datasets

Anonymised WSIs of human medical liver biopsies from N=545 patients were gathered retrospectively from the archives at Leeds Teaching Hospitals NHS Trust from 2016-2022 (Table 2). Image data was obtained from the UK National Pathology Imaging Co-operative after ethical approval from the Office for Research Ethics Committees Northern Ireland (ORECNI), research ethics committee reference: 22/NI/0033. Eligibility criteria for inclusion were adult liver cases with diagnosed MASLD. Each case includes several biopsies stained with Van Gieson (VG), Haematoxylin and Eosin (H&E), or both stains. Kleiner scores were automatically extracted from clinical text reports using a Python script. To achieve this, all text reports with instances of the term ‘Kleiner’ were identified (N=149). Next, appropriate regular expressions were applied to retrieve semiquantitative scores for steatosis (N=125) and fibrosis stage (N=149). For manual verification of the extracted scores, the paragraphs containing the term ‘Kleiner’ were also exported to an Excel spreadsheet. From those cases with known fibrosis stage, a total of 136 VG-stained WSIs were available for evaluation of fibrosis quantification. From those cases with known Kleiner scores for steatosis, a total of 115 H&E WSIs were available. Since Kleiner scores were reported for cases with mild, moderate, or severe steatosis, we further searched reports for cases with ‘no steatosis’ (N=91) resulting in a total of 206 H&E WSIs for evaluation of the proposed steatosis quantification method. Image quality assessment was performed manually over the slides to exclude out-of-focus or blur samples. One H&E slide was identified with a partial out-of-focus issue in some regions.

**Table 2.**
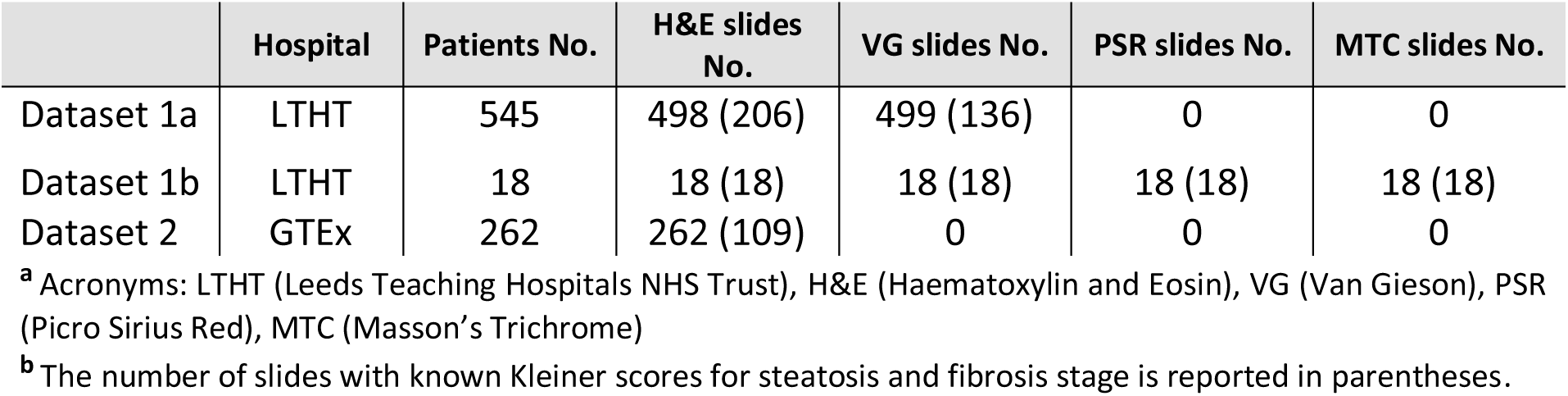
Dataset Summary.

To further study the effects of different dyes on fibrosis quantification, N=18 resection cases with background MASLD were identified ensuring representation of all stages of fibrosis (Dataset 1b, Table 2). Samples from each case were then stained with PSR, MTC, and VG. Image quality assessment was performed manually over these slides and one MTC slide with fibrosis stage of 0 was excluded due to severe staining artifact.

We further validated our steatosis quantification algorithm on an external public dataset using H&E WSIs from the Genotype-Tissue Expression (GTEx) public portal (Broad Institute, Cambridge, MA, USA) [33]. The GTEx tissue image repository includes a diverse collection of histology images derived from various tissue types sourced from postmortem donors. For this study, we selected N=262 liver biopsy cases exhibiting steatosis. Kleiner scores were automatically extracted from reported pathology notes for N=109 cases using a Python script. To achieve this, pathology notes were scanned for the steatosis proportionate area (SPA) and then categorised in four groups: 0 (SPA≤ 5), 1 (5 < SPA ≤ 33), 2 ( 33 < SPA ≤ 66), and 3 (66 < SPA) (Table 2).

### 2.3. Liver Quantification

The overall workflow to quantify steatosis and fibrosis in WSIs includes three main steps. First, the foreground is segmented using colour thresholding and morphological operations. Second, each WSI is divided into small tiles, e.g. 2048×2048 pixels. Each tile is then processed independently and, finally, results are aggregated from all tiles into a single score.

### 2.4. Foreground Segmentation

In digital pathology WSIs, the background glass slide is often white but may contain ink markings or other types of artefacts. In the absence of artefacts, the white background can be segmented using colour thresholding in the Hue-Saturation-Value (HSV) space; pixels with saturation below 4% and a value above 90% were heuristically selected as background in this study. The foreground was then derived by negating the background mask. This approach may result in segmenting non-white artefacts as foreground. To avoid these artefacts, tissue can be directly segmented by using appropriate thresholds. In this study, we set these thresholds heuristically for each stain independently: For H&E staining, the hue range was [240, 360], the saturation range was [4, 100], and the value range was [30, 100]. For VG staining, the hue range was [-60, 60] and the saturation and value were similar to H&E staining. Morphological operations were then applied to refine the segmented regions and remove noise or small artefacts.

### 2.5. Steatosis Quantification

Fig. 1 provides an overview of the fat quantification workflow used in this study. First, white regions are segmented using a pixel-thresholding method in the HSV-space; each pixel with saturation below 10% and a value above 70% was segmented. These thresholds were heuristically set in this study. For each segmented region, contours were extracted to estimate three morphological features: solidity, elongation, and diameter of the minimum enclosing circle. Based on these metrics, each white region is classified as fat (isolated globules), unknown (potentially containing overlapping fat globules), or other non-fat structures. The unknown group is further segmented into smaller regions using watershed segmentation to separate overlapped fat globules. Each white region is then classified as fat or other non-fat structures. All white regions identified as fat are then combined into a single mask. Finally, SPA is computed as the total area occupied by fat globules divided by the segmented tissue area. By construction, SPA has a value between 0 and 1.

Using visual assessment on five image tile samples by a consultant pathologist, we heuristically set thresholds for each morphological feature as follows: for a fat globule, solidity was in the range [0.85, 1], elongation was in the range [0.4, 1], and the diameter was in the range [5, 100] micrometres. For unknown white regions comprising overlapping fat globules or non-fat regions, solidity was in the range [0.7, 1], the elongation was in the range [0.05, 1], and the diameter was greater than 5 micrometres. Other remaining white regions were excluded as non-fat structures.

To avoid double-counting identified fat globules, the WSI is divided into overlapping tiles. The amount of overlap must be greater than the maximum fat globule radius present in the image (Supplementary Fig. 2). Those globules with centres inside the overlapped region are excluded from the current tile and are counted in the neighbouring tiles.

### 2.6. Fibrosis Quantification

For fibrosis quantification, tissues stained with PSR, MTC, and VG were used in this study. To identify collagen, colour segmentation is performed using pixel thresholding in the HSV space. The hue channel can efficiently discriminate between collagen and liver tissue (Fig. 2). Routine variation in slide staining and image capture processes requires automated solutions to incorporate flexibility in the form of image standardisation and/or adaptive thresholding. Here, we studied both approaches. First, we employed the stain normalization algorithm developed by Macenko et al. [34] to eliminate variations in colour profiles and standardise the stain appearance across all slides in the dataset [Fig. 2]. Next, to identify the optimal threshold for the hue channel, a Gaussian mixture model with 2 components was fitted to the histogram of hue values in the tissue and the maximum likelihood threshold was estimated similar to Masseroli et al. [28]. Further, following the segmentation of fibrotic tissue, CPA is computed as the proportion of the total collagen area to the segmented tissue area. By definition, CPA holds a value between 0 and 1.

### 2.7. Evaluation

To assess the performance of the proposed method, several evaluation metrics are employed. The Spearman rank coefficient (*ρ*) is calculated to measure the correlation between the computed CPA/SPA values and the Kleiner scores. For visual evaluation, Box-and-whisker plots are presented to provide a graphical representation of the data. To measure the reproducibility of SPA, the coefficient of variation root mean squared error (CV-RMSE) was computed using triple measurements for each H&E biopsy sample:

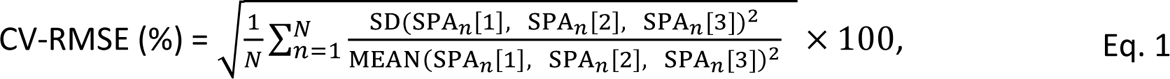

Where SPA_*n*_[*i*] is the SPA value for case *n* and tissue level *i* ∈ [1, 2, 3], and SD is the standard variation of the measurements. To measure consistency between CPA measurements across three different staining methods, Bland-Altman plots were used in this study.

## 3. Results

### 3.1. Background Segmentation

All WSIs (N=997) were segmented using the proposed colour thresholding algorithm. The quality of segmented maps was visually assessed and N=89 maps were manually corrected using brush tools in QuPath version 0.5.1 [35]. Supplementary Figure 2 shows two examples where colour thresholding failed, and foreground masks were manually corrected.

### 3.2. Steatosis Quantification

Each WSI in dataset 1a (Table 2) included three tissue sections of the core needle biopsy sampled at different levels in the paraffin block. To assess the reproducibility of SPA, CV-RMSE was computed using triple measurements for each biopsy sample, one on each of the three different sections taken of the tissue at different levels. The estimated CV-RMSE in this dataset measured at 40x magnification was 13.7%.

A strong correlation (*ρ*=0.92) was observed between SPA and the Kleiner scores at 40x magnification using dataset 1a (Table 3). Fig. 3(A) shows the box plot for visualising the distribution of SPA per each Kleiner score. Fig 3(B) shows a similar analysis for the estimation of the proportionate area of white regions. The highest range for SPA was approximately 25% compared to 40% for the proportionate area of white regions. Notably, these observations suggest that pathologists tend to overestimate the amount of fat with values exceeding 66% for cases with a Kleiner score of 3. Almost no overlap between Kleiner scores 0 and 1 was observed for SPA (Fig. 3A) whereas substantial overlap existed for white regions proportionate area (Fig. 3B). This finding highlights the efficacy of morphological filters to exclude non-fat white regions. Overall, SPA provides better discrimination between Kleiner scores compared to white regions proportionate area. Fig. 3C shows the box-and-whisker plot between estimated SPA and Kleiner scores using the public external dataset 2. The distribution of SPA per each Kleiner score was similar to the pattern observed in dataset 1a (Fig. 3A). However, the Spearman rank correlation was lower in the second dataset.

**Figure 3.**
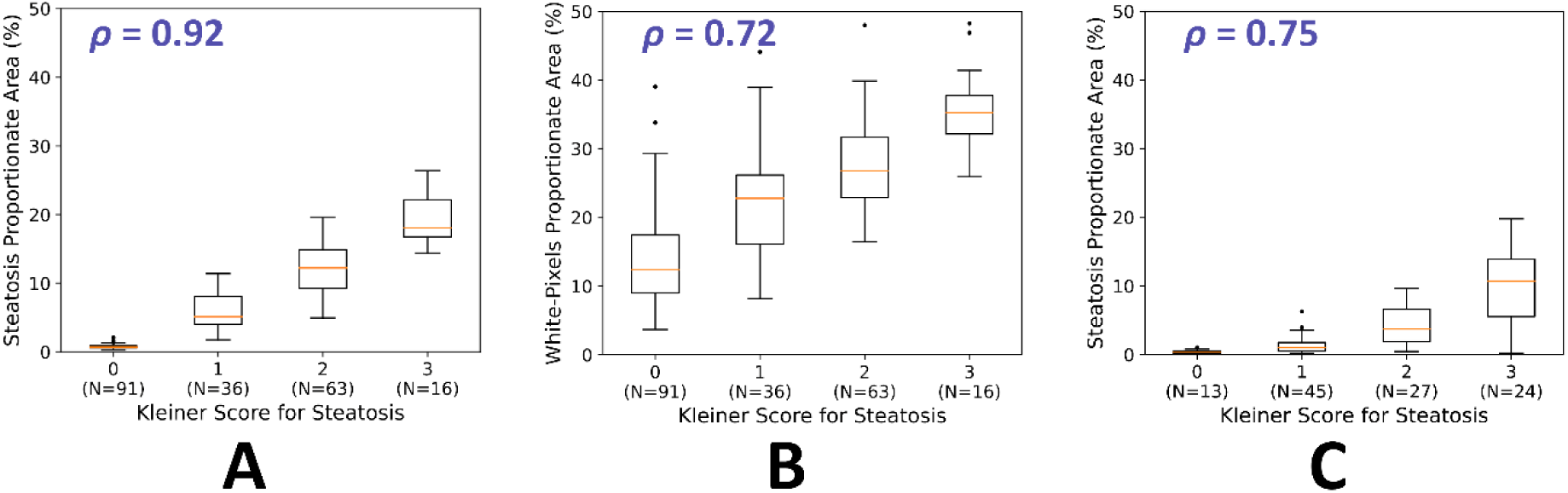
A) Box-and-whisker plots illustrating the distribution of estimated steatosis proportionate area (SPA) in relation to Kleiner scores. A strong correlation (ρ=0.92) was identified between SPA measurements at 40x magnification and Kleiner scores within a cohort of N=206 cases from dataset 1a. For each subject, three tissue-level cuts were analysed, and the maximum proportionate area was reported. B) Box-and-whisker plots depicting the relationship between estimated white pixel proportionate area and Kleiner scores. A moderate correlation (ρ=0.72) was observed within a cohort of N=206 cases from dataset 1a. Excluding non-fat white regions significantly improves the separation between different Kleiner scores as shown in panel A. C) Box-and-whisker plots between estimated SPA at 20x magnification and Kleiner scores on the external public dataset 2. A moderate correlation (ρ=0.75) was observed in a cohort of N=109 cases.

**Table 3.** The impact of magnification on steatosis quantification.

|  | Steatosis Quantification |  | White Pixel Counting |  |
| --- | --- | --- | --- | --- |
| | Time [sec] | $\rho$ | Time [sec] | $\rho$ |
| 40x | 195 | 0.92 | 38 | 0.72 |
| 20x | 112 | 0.91 | 47 | 0.72 |
| 10x | 71 | 0.90 | 35 | 0.63 |
| 5x | 27 | 0.63 | 4 | 0.43 |

\* $\rho$ = Spearman Rank Correlation Index between estimated proportionate area and Kleiner scores for steatosis

Table 3 shows the effect of magnification on the algorithm in terms of run time and accuracy using dataset 1a. Employing a lower magnification of 10x can speed up the code by a factor of 3 compared to 40x magnification while maintaining the performance in terms of the Spearman rank correlation index (*ρ* ≥ 0.9). However, the mean value for SPA decreased sharply with respect to magnification levels as shown in Fig. 7A suggesting 40x or 20x magnification is essential for accurate SPA measurements.

### 3.3. Fibrosis Quantification

A moderate correlation (*ρ* = 0.51) was observed between estimated CPA and provided semi-quantitative fibrosis stages by pathologists (Fig. 4). Fig. 4 shows the box plot for CPA estimation at 1.25x magnification. The median CPA was 8.2%, 10.7%, 13.2%, 15.8%, and 20.3% for fibrosis stages 0 to 4, respectively.

**Figure 4.**
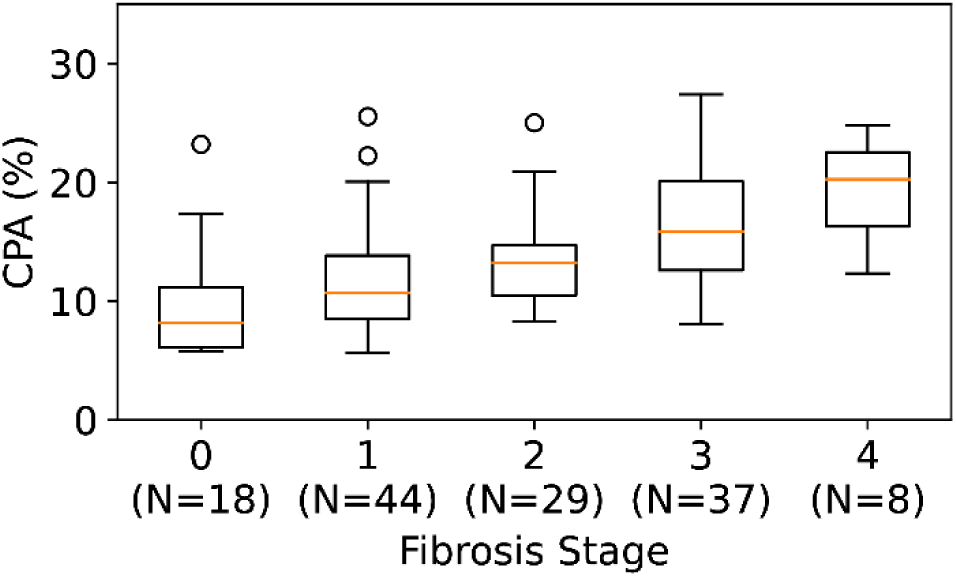
Box-and-whisker plots for the distribution of estimated collagen proportionate area (CPA) with respect to Kleiner scores in the first dataset with VG staining (N=136). The Spearman rank coefficient index (*ρ*) was 0.51.

To further study the effects of different dyes on CPA estimation, N=18 resection cases of MASLD were identified and stained with PSR, MTC, and VG. Fig. 5 shows box plots for estimated CPA at 1.25x magnification. The proposed method (last row in Fig. 5) includes stain normalisation followed by adaptive thresholding. A strong correlation (*ρ* ≥ 0.85) was observed between CPA and the fibrosis stages using all three stains. A huge overlap in CPA was observed across stages 0 to 3 making it difficult to distinguish between these stages. However, a clear cut-off threshold of CPA=13% was observed to separate cases with fibrosis stages 3 or 4 from the other cases. To assess the variation between each stain pair, Bland-Altman plots were used in this study. Figure 6 suggests no significant bias (less than 1%) between PSR and MTC in the measurement of CPA. However, CPA measurements using MTC and PSR were on average 3% higher than the corresponding measurement using VG. Overall, an excellent agreement was observed among the three stains.

**Figure 5.**
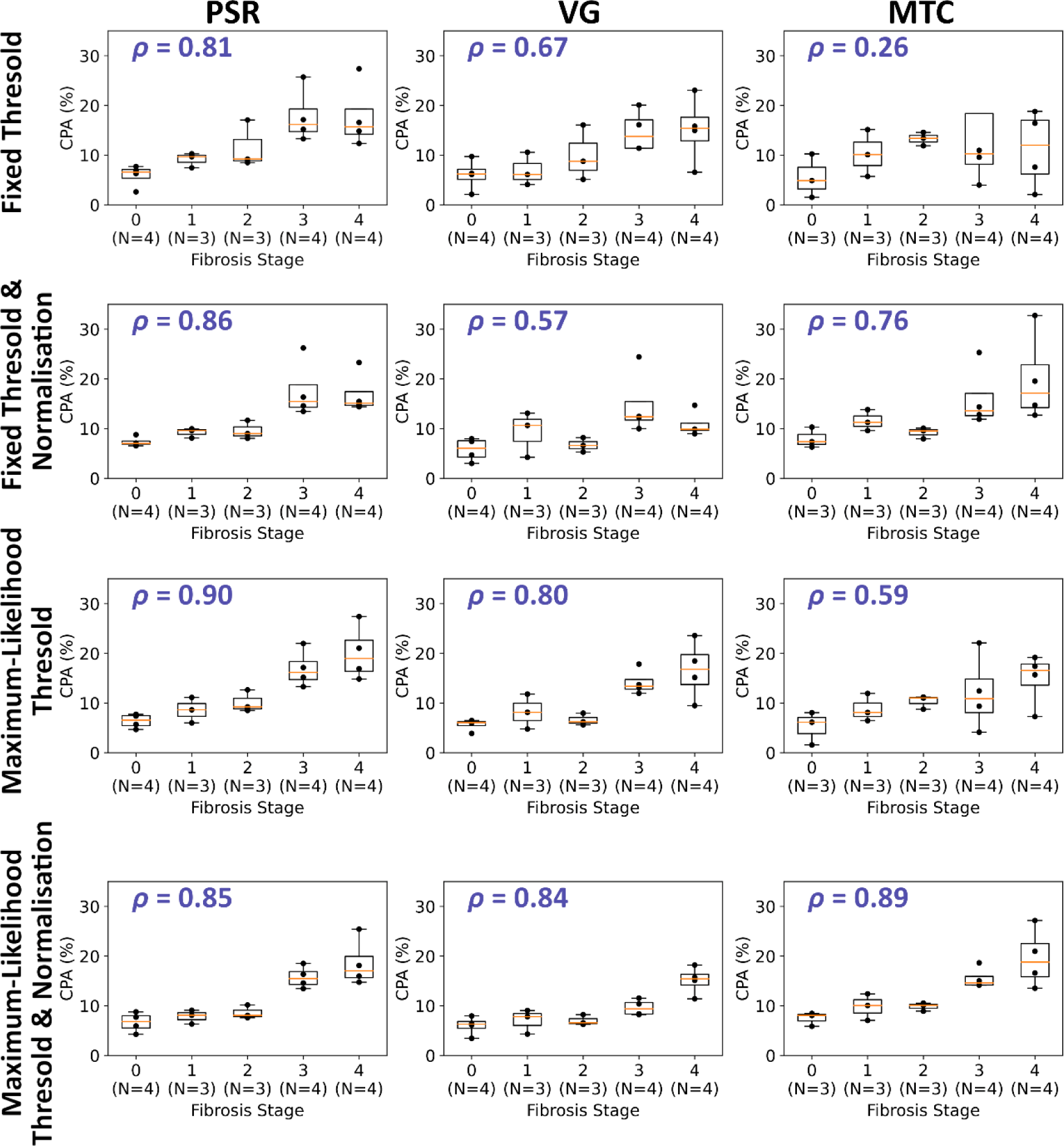
Box-and-whisker plots for the distribution of estimated collagen proportionate area (CPA) with respect to Kleiner scores in the second dataset (N=18). Each column, from left to right, shows results for Picro Sirius Red (PSR), Van Gieson (VG), and Masson’s Trichrome (MTC) staining methods, respectively. Each row shows a different quantification method. The Spearman rank coefficient index (*ρ*) is reported for each plot separately. Results are reported at 1x magnification.

**Figure 6.**
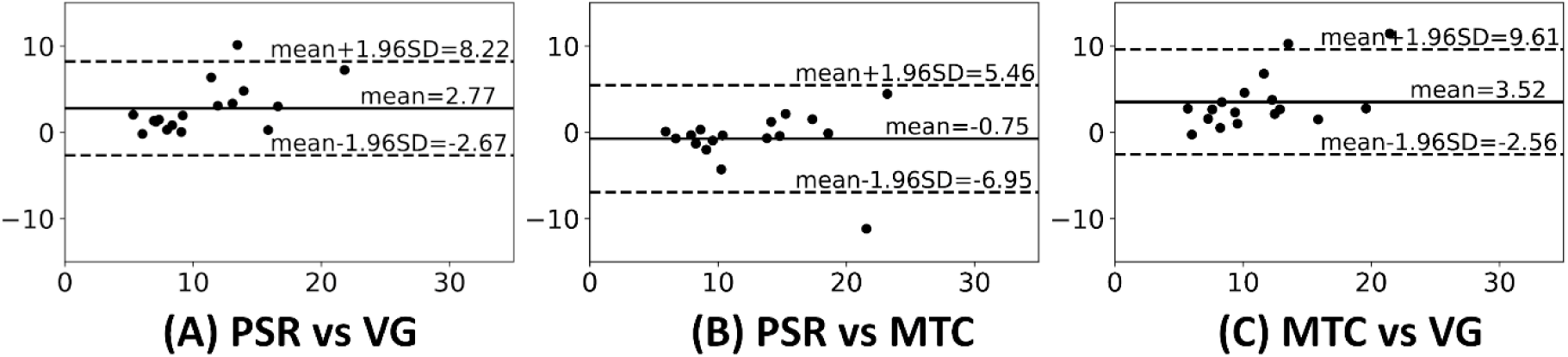
Bland-Altman plots for comparing estimated collagen proportionate area (CPA) between each staining pair. The *x*-axis shows the mean CPA in percent and the *y*-axis shows the difference between paired CPA measurements.

To reliably segment fibrotic tissue both stain normalisation and maximum likelihood adaptive thresholding were employed in this study. To evaluate how each step contributes to the CPA estimation, we excluded either the normalisation step or the adaptive thresholding. The three variations of the proposed method were visualised in rows 1 to 3 in Fig. 5. Among the three staining dyes used in this study, only PSR had an acceptable performance (*ρ* ≥ 0.8) in all experiments. This finding might suggest PSR as the gold standard for quantitation of fibrosis. In the absence of adaptive thresholding, stain normalisation boosted the CPA performance most in MTC staining by increasing the Spearman rank correlation from 0.3 to 0.8. In the absence of stain normalisation, adaptive thresholding improved the CPA performance in all stains.

To assess the effect of magnification on fibrosis quantification, we performed CPA estimation at different magnifications, i.e., 40x, 20x, 10x, 5x, 2.5x, and 1x, using the PSR stain in the second dataset and reported the Spearman rank correlation index (*ρ*) and run time in Table 4. The CPA estimation tends to increase on average by 2% for fibrosis stages 0 to 3 by increasing the magnification level from 1.25x to 40x. However, the CPA reduced on average by 3% for fibrosis stage 4 when increasing the magnification level from 1.25x to 40x using the PSR stain (Fig. 7B).

**Figure 7.**
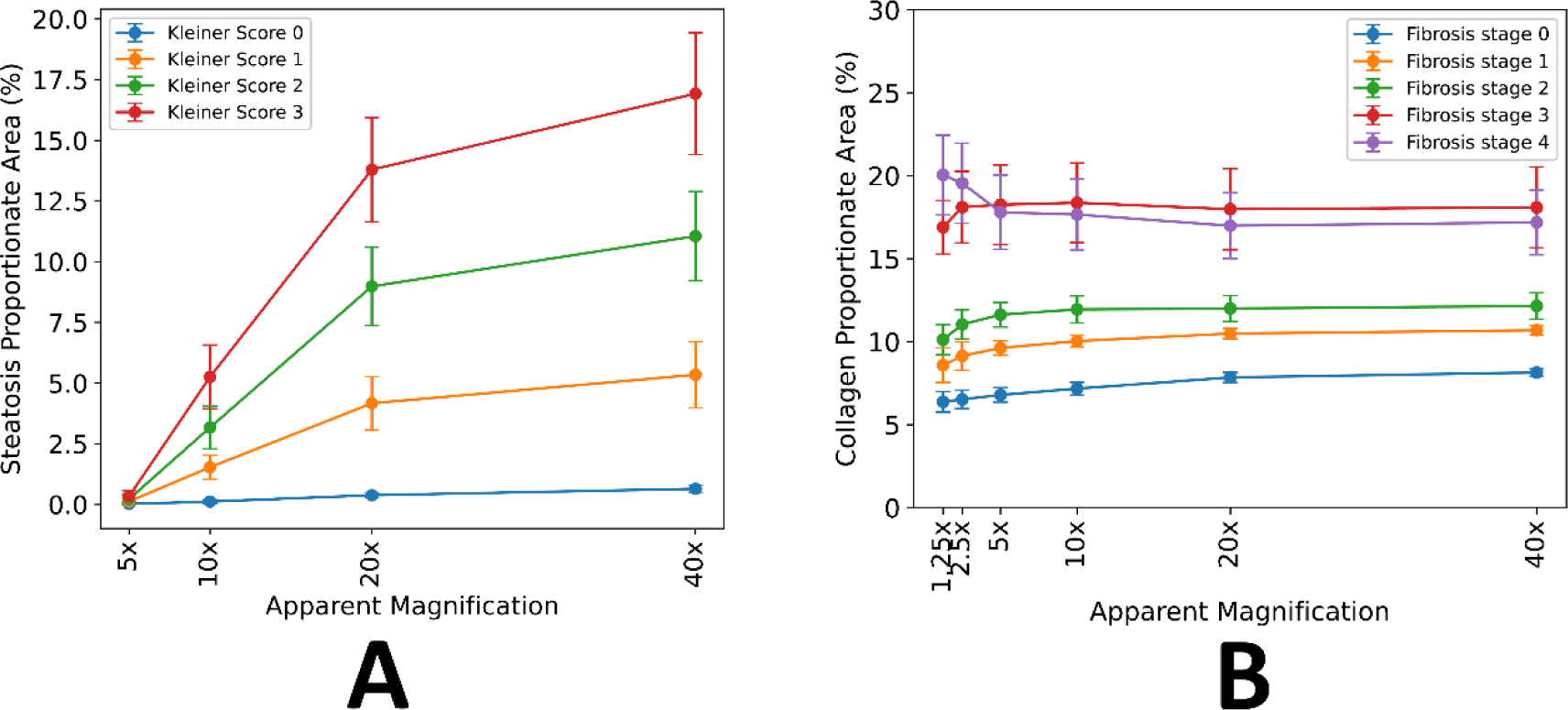
Magnification effect on Steatosis Proportionate area (SPA) and collagen proportionate area (CPA). Filled-in circles represent the mean values, and the error bars show 0.5 standard deviations (SD). Scaling SDs was to improve the visual representation of results. (A) SPA reduces sharply as apparent magnification decreases from 40x to 5x. The lowest magnification to preserve the separation between classes was 10x but for accurate estimation of SPA, at least 20x magnification is required. (B) Increasing the apparent magnification only modestly increases the CPA values except for cases with a fibrosis stage of 4. At higher magnification, separation between stage 3 and 4 is more challenging but overall the CPA correlation with fibrosis stage remains the same (Table 4).

**Table 4:** The impact of magnification on Collagen Proportionate Area (CPA) estimation using the PSR staining technique.

| | Time [sec] | $\rho$ |
| --- | --- | --- |
| 40x | 774 | 0.88 |
| 20x | 529 | 0.88 |
| 10x | 79 | 0.87 |
| 5x | 59 | 0.88 |
| 2.5x | 7 | 0.89 |
| 1x | 2 | 0.90 |

We further compared our method against state-of-the-art AI based quantification method proposed by Heinemann *et al.* [16] using N=18 cases stained with MTC. Note that their method was trained on data stained with MTC only and did not work on PSR and VG stains. Figure 8 shows box plots for the distribution of AI fibrosis scores and measured CPA values. Their performance in terms of correlation with Kleiner scores were similar. A mild correlation (*r*^2^ = 0.58) was observed between the AI fibrosis scores and the corresponding CPA measurements.

**Figure 8.**
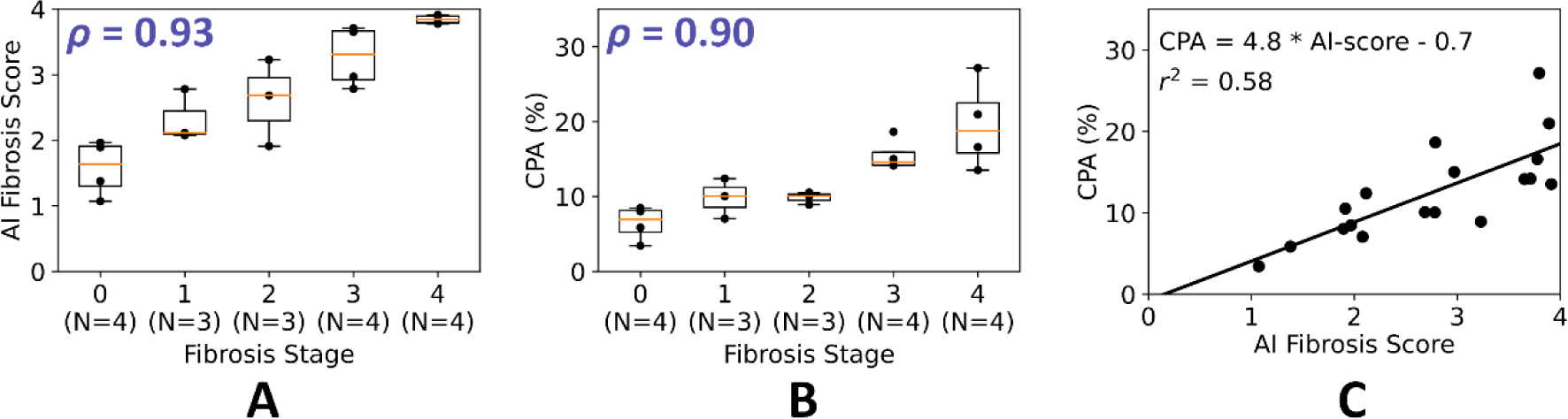
(A) Box-and-whisker plots for the distribution of estimated AI fibrosis score [16] with respect to Kleiner scores in the second dataset stained with Masson’s Trichrome (MTC). The method proposed by Heinemann *et al.* [16] is based on deep convolutional neural networks and produced scores in the same range as Kleiner scores. (B) Box-and-whisker plots for the distribution of estimated collagen proportionate area (CPA) with respect to Kleiner scores in the second dataset stained with MTC. (C) A moderate correlation (*r*^2^ = 0.58) was observed between estimated CPA using classical image analysis and the AI fibrosis score proposed by Heinemann *et al.* [16]. Note the AI method is applied as published by authors without any further training on our dataset.

## 4. Discussion

This paper presents an open-source Python package for the quantification of steatosis and fibrosis in MASLD. The proposed algorithm does not require training using manual annotations and only relies on colour and morphological features to segment regions attributed to fat globules and fibrosis. We reported the proportionate area of segmented features in relation to the total tissue area as SPA and CPA, respectively, in a large evaluation dataset of 424 cases. We further validated the steatosis quantification algorithm on a public external dataset with 109 cases.

The proposed steatosis quantification algorithm can improve quantitative tissue characterisation over human pathological evaluation by increasing both reproducibility and accuracy. The estimated CV-RMSE across N=205 cases, each containing three tissue levels, was 13.7%. A strong correlation (*ρ* = 0.92) was observed between estimated SPA and steatosis Kleiner scores. The highest SPA observed in this study was approximately 25%, which was in a sample with a Kleiner score of 3. This observation suggests that pathologists tend to overestimate the amount of fat in histology images with SPA values exceeding 66% for a Kleiner score of 3 [36]. Although the score is meant to be an estimate of the number of hepatocytes containing fat, visual assessment of this feature by pathologists is difficult and in practice many are probably estimating the surface area of the liver involved by steatosis – analogous to a calculated SPA. This finding of human overestimation of SPA is in keeping with results reported in previous research [9], [16]. The estimated correlation index in this study was comparable to the state-of-the-art methods employing deep convolutional neural networks for fat quantification (c.f. Table 1). This finding may suggest promoting classical image analysis for routine clinical use given its low complexity compared to deep learning methods.

In this work, we employed morphological filters to identify fat globules. To assess the contribution of these filters, we compared the measured SPA versus the proportionate area of white regions. While excluding the morphological filters accelerated the algorithm by a factor of 5, the performance dropped notably as measured by the Spearman rank correlation index (*ρ* = 0.92 for fat detection compared to *ρ* = 0.72 for white pixel counting).

The proposed fibrosis quantification algorithm resulted in a moderate correlation (*ρ* = 0.5) between the measured CPA and the fibrosis stages using 136 slides stained with VG. The estimated correlation index in this study was comparable to the state-of-the-art methods employing deep convolutional neural networks (Table 1). However, employing a smaller dataset (N=18) ensuring a uniform representation of all stages of fibrosis resulted in higher correlations (*ρ* = 8). The CPA biomarker has several advantages over semiquantitative fibrosis scores including high reproducibility and increased sensitivity in detecting changes in hepatic fibrosis [31]. However, CPA has a few limitations. First, CPA does not reflect the architectural pattern or type of fibrosis distribution, limiting its ability to provide detailed information about liver fibrosis characteristics or their pathophysiological impact. Second, there is a significant overlap in CPA values across different semi-quantitative fibrosis stages, which can lead to challenges in accurately classifying fibrosis severity based solely on CPA measurements.

We studied the impact of image magnification on quantification results. Overall, computation cost is reduced at lower magnifications allowing for fast quantifications. In steatosis quantification, estimated SPA reduces sharply when reducing the magnification levels from 40x to 10x on average by 10%. Table 3 suggests 10x as the optimum magnification for steatosis quantification considering run time and correlation with Kleiner scores. However, 40x or 20x magnification levels were found to be essential for accurate SPA measurements (Fig. 7A). In fibrosis quantification, magnification levels had a modest effect on CPA measurements and lower magnifications up to 1x were found to be well correlated with pathologist scores (Table 4 and Fig. 7B).

We studied three different stains commonly used in clinical practice for fibrosis quantification. No systematic difference was observed between PSR and MTC in CPA measurement (bias=-0.7%, p-value=0.4), but both PSR and MTC stains resulted in slightly higher CPA values compared to VG on average by 3% (p-value < 0.001) (Fig. 6). Using the proposed stain normalisation and maximum likelihood thresholding in the HSV space, all three stains had a similar performance in terms of correlation with pathologist scores. However, the PSR stain had consistently better performance using different quantification algorithms (Fig. 5).

This study had a few limitations. First, Kleiner scores were obtained from a single observer limiting our ability to assess inter-observer variability for the quantification task. It would be advantageous to involve multiple observers in future research to enhance the robustness of our findings by reporting correlations based on the average scores.

Machine learning-based and image analysis tools can improve quantitative tissue characterisation upon human pathological evaluation by increasing reproducibility, identifying features associated with clinical outcomes, and providing a platform for rigorous and consistent assessment of disease regression following treatment [11]. In this study, we developed an open-source tool to quantitate two pathological features of interest in the MASLD. Our results were promising but further evaluation using larger dataset from different laboratories may be warranted in future.

## Data Availability

All data [except data collected at Leeds Teaching Hospitals NHS Trust] are available online at https://github.com/mfarzi/liverquant and https://gtexportal.org/home/histologyPage

## Additional Information

Liver-Quant package can be downloaded from https://github.com/mfarzi/liverquant. The first dataset used in this study comprising slides from Leeds Teaching Hospitals NHS trust will be published as part of the Big-Picture project (see https://bigpicture.eu/). The second dataset used in this study is publicly available from https://gtexportal.org/home/histologyPage.

## Supplementary

**Supplementary Figure 1.**
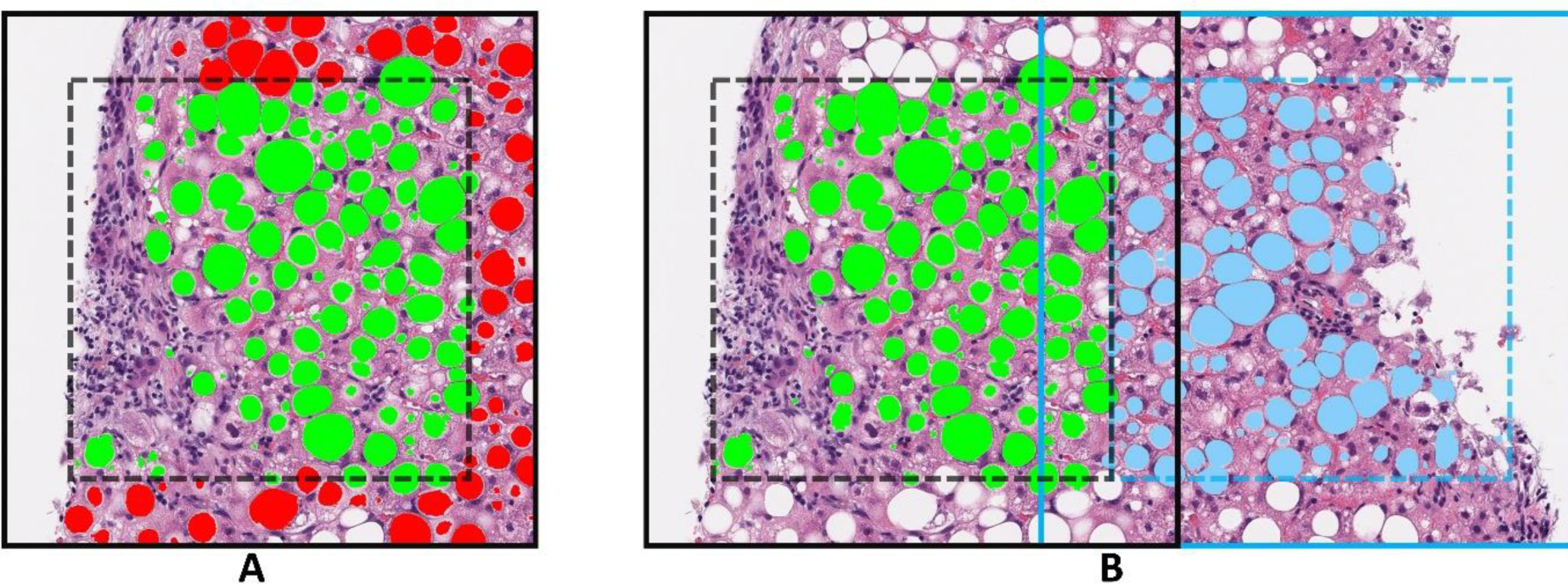
A) The solid line shows the image tile and the dashed line shows the overlapped regions. All globules with their centres lying in the dashed square are marked as green and counted in this tile. The globules marked in red with their centres lying in the overlapped region are excluded. **B)** Two image tiles are shown using solid black and blue lines. Globules detected per each tile are marked in green and blue, respectively.

**Supplementary Figure 2.**
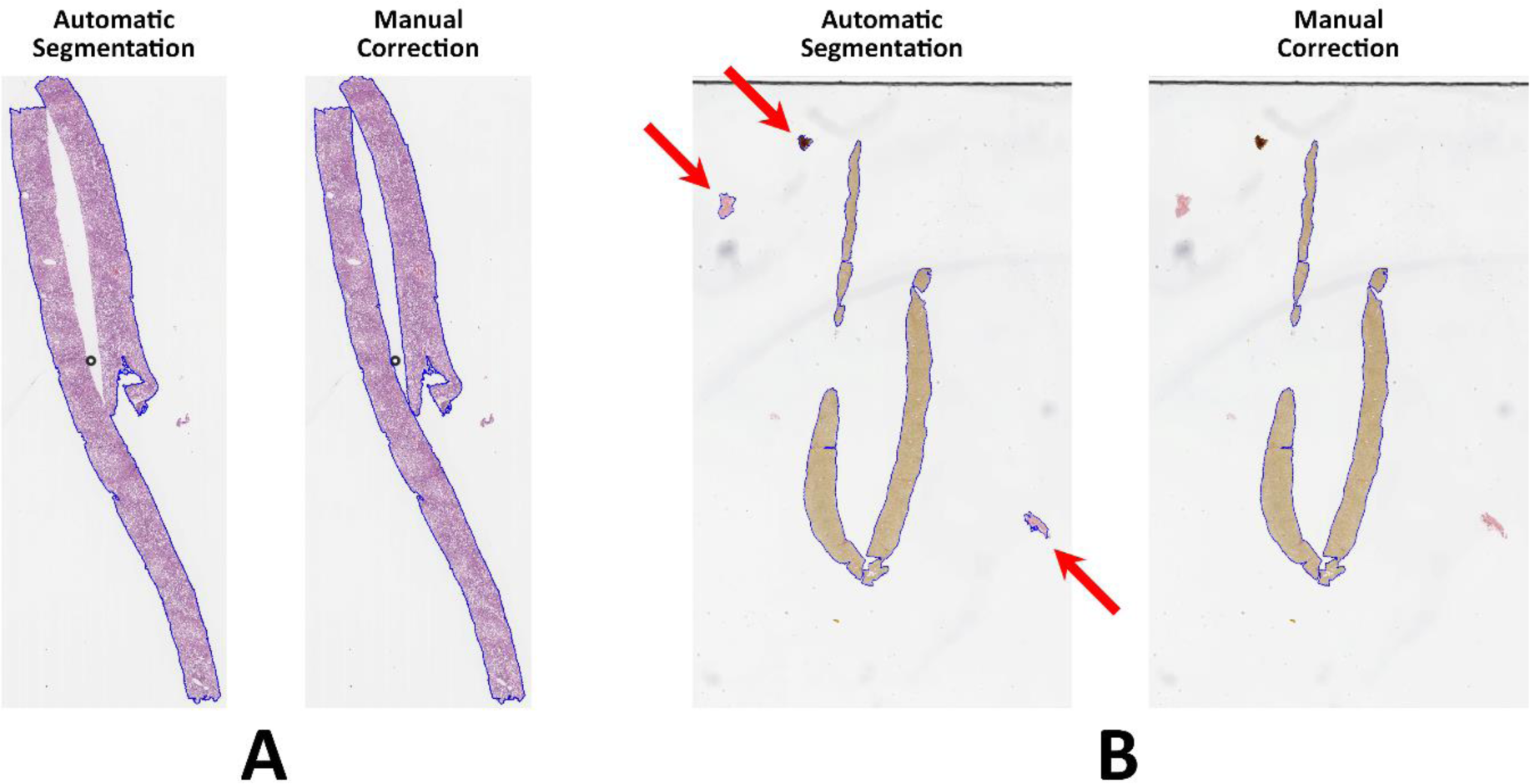
Two examples of failed automatic foreground segmentation using colour thresholding in the HSV-space. The foreground is marked with blue contours. A) Automatic segmentation for an H&E slide resulted in a hole at the intersection of two biopsies. The hole was removed manually using QuPath software. (B) Automatic segmentation of a VG slide was failed due to the presence of artefacts in the background identified by red arrows. The contours were removed manually using the QuPath Software.

